# Elective genomic sequencing for adults in research, clinical and commercial contexts

**DOI:** 10.64898/2026.06.09.26355296

**Authors:** Michael D. Linderman, Sophia M. Adelson, Tala M. Berro, Jennifer L. Anderson, Scott D. Crawford, Tshaka J. Cunningham, Edward D. Esplin, Altovise T. Ewing-Crawford, Daiva E. Nielsen, Stacey Pereira, Tara Schmidlen, Heather Andrighetti, Steven B. Bleyl, George M. Church, Eden V Haverfield, Madhuri Hegde, Lazaridis N. Konstantinos, Paul Kruszka, Debra Leonard, Thomas May, Molly McGinniss, Vaibhav Pandya, Eric E. Schadt, Bastian Greshake Tzovaras, Bethany Zettler, Amy L. McGuire, Robert C. Green, the PeopleSeq Study Team

**Author notes:** Corresponding Authors: Michael Linderman, Robert Green.

## Abstract

**Purpose:** Elective genomic sequencing (EGS) returns monogenic disease findings in multiple genes, including potentially novel variants, and may also provide participants with carrier status, pharmacogenomic and other health-related information. The PeopleSeq Study assessed participants’ motivations for and concerns about EGS and the associated clinical and psychosocial outcomes across diverse EGS providers.

**Methods:** We administered a shared questionnaire to participants who chose to undergo EGS via 18 academic, clinical, or commercial EGS platforms.

**Results:** We enrolled 1575 participants, of whom 1147 (72.8%) completed a questionnaire after receiving their EGS results. A majority (60.3%) of the participants who completed a post-result questionnaire self-reported receiving results they assessed as important, including negative findings, and 75.9% reported a form of health-related utility. Among a subset (19.4%) who shared their EGS reports, 16.6% (37 of *n*=223) received a monogenic finding and self-reported results deemed “important” were consistent with EGS reports. Most participants (74.1%) discussed their results with their family, but fewer discussed their results with a healthcare provider other than the site team (41.7%) or had one or more medical visits as a direct result of their EGS testing (23.1%). Participants expressed diverse motivations for EGS, with 91.4% expressing interest in their personal disease risk and 54% who expressed quasi-indication-based motivations related to family medical history. Individuals motivated by family history reported important results at a significantly higher rate.

**Conclusions:** Early adopters of EGS are motivated by general interest in their health as well as quasi-indication-based considerations such as family history. A majority of participants learned results they considered medically important, but a much smaller segment engaged healthcare providers with their results.

## Introduction

Genomic sequencing is widely used for rare disease diagnosis and treatment, and personalized cancer treatment, and now is increasingly used for screening purposes in ostensibly healthy individuals[1, 2]. Elective genomic sequencing (EGS), also described as “predispositional personal genome sequencing” or “clinical genome screening”, is now available in many forms as an elective clinical test[3]. Here we define EGS as sequencing-based testing that returns monogenic disease findings in multiple genes to participants, including potentially novel variants (distinguishing EGS from array-based direct-to-consumer (DTC) genotyping). Depending on the test implementation, EGS results may include a varying number of genes, carrier status to inform reproductive planning, pharmacogenomic data to guide current and future medication choices, and risk information for polygenic conditions[1]. The goal of EGS is to report genetic findings that provide risk stratification and inform participants’ future medical management.

In 2021 the American College of Medical Genetics and Genomics (ACMG) published a points to consider (PTC) statement about the potential opportunities and challenges for EGS[3]. The PTC highlighted the need for additional data about the use of this testing, especially outside traditional genetic health-care delivery models. Several research projects have studied the clinical and personal outcomes of EGS[4–13]. However, these projects were generally small and conducted as controlled research protocols in academic settings, a different context than the clinician-ordered testing that is now available. Large-scale clinical testing has provided new insights into rates of positive findings, carrier results, and cascade testing[14–17], but those laboratories are often not able to collect data about patients’ motivations, clinical and personal utility, and psychological outcomes in a systematic way. Thus, although thousands of individuals are undergoing EGS, little is known about the personal motivations and outcomes of this testing as it is implemented more broadly.

The Personal Genome Sequencing Outcomes (PeopleSeq) Consortium is an NIH-funded collaborative effort by 18 academic, clinical, and commercial EGS projects/laboratories. PeopleSeq investigators designed questionnaires and collected survey data on individuals who chose to undergo EGS. PeopleSeq questionnaires assessed participants’ motivations and concerns about testing, their resulting genetic findings, participants’ perception of the importance and utility of those results, and their reports of the associated clinical and psychosocial outcomes. The different collaborating projects represent the breadth of contexts in which EGS is in use. Here we present observational findings on participants’ motivations for testing, genetic findings, and clinical outcomes after receiving their genetic results.

## Methods

### Overview

The PeopleSeq Consortium[18] included 18 EGS sites/projects (see Supplemental Table 1 for an overview of each site and Linderman et al. for a more detailed review of these and other similar projects[1]). Collaborating sites included academic research projects, industry laboratories that offered direct access to EGS testing, clinicians that order EGS tests from one or more laboratories and “third-party” sites where individuals could share their own genetic data. The specific test scope (e.g., panel vs. ES vs. GS), cost to the participant, pre-test counseling, results provided and disclosure protocols differed between sites and are listed in Supplemental Table 1. Although the scope of EGS results differed between projects, all tests reported monogenic disease findings, including novel deleterious variants that qualified as likely pathogenic variants, in numerous genes.

PeopleSeq Consortium investigators conducted the PeopleSeq Study, which administered web-based questionnaires to participants at each collaborating site. Participants were recruited through heterogeneous, non-probability pathways. As a result, the sample does not represent a single defined sampling frame, but instead a cross-section of individuals engaging with EGS in academic, clinical, and commercial settings. Consent for participation in the PeopleSeq Study was separate from, and in addition to, consent to undergo EGS at the collaborating site. The study itself did not perform any genetic testing or disclose results to participants, but did, through a supplemental award to increase the cohort diversity, fully fund EGS via Genome Medical for 49 participants recruited via educational events organized with primarily African American community groups. Funded individuals were invited to participate in the PeopleSeq study but were not required to do so to undergo testing.

Participants were invited to enroll into the PeopleSeq Study at different points in the testing process depending on the workflow at each site. The study workflows are summarized in Figure 1. If a site was conducting EGS on an ongoing basis and could contact participants prior to results disclosure, individuals would be invited to complete a pre-disclosure (“pre”) questionnaire after deciding to proceed with testing but before they received their results and a second post-disclosure questionnaire (“post”) after receiving their results. If participants could not be contacted prior to results disclosure (e.g., because of that site’s workflow, or results had already been disclosed when the study began recruiting) participants were invited to a “catch-up” questionnaire that included all the pre- and post-disclosure items (both groups received the same items). The post or catch-up questionnaire was administered a minimum of 8 weeks after anticipated results disclosure (as reported by the collaborating site). All participants received an annual follow-up questionnaire. The Mass General Brigham (previously Partners) HealthCare Human Research Committee Institutional Review Board approved the study. Each site consulted their own Institutional Review Board, or legal team, as applicable and received additional approval if necessary.

**Figure 1:**
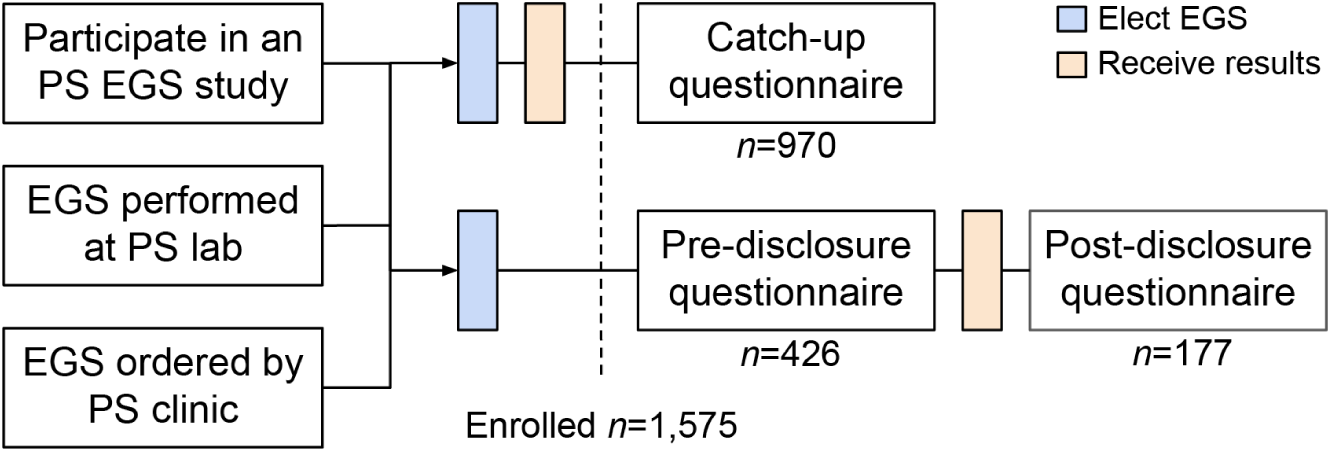
Study flowchart. Participants accessed EGS via research studies, clinics and laboratories collaborating in the PeopleSeq (PS) consortium. All participants were enrolled after electing EGS. Participants who had already received their results received a single “catch-up” questionnaire; if participants had not yet received their results, they received a “pre-disclosure” survey before and “post-disclosure” survey after receiving their results. All sites used the “catch-up” workflow; Harvard PGP, Illumina UYG, UVM, MedCan, Genome Medical and GeneDx also used the pre/post workflow. See Supplemental Table 1 for site details. The total number of potential eligible and invited individuals is estimated to be 6,750. A majority of the eligible and invited participants (6,431) could be tracked, but a fraction of cohorts recruited participants through passive advertisements (viewed by an unknown number of individuals) or e-mail announcements sent to an unknown number of individuals.

### Survey design and measures

The PeopleSeq questionnaires were developed by an interdisciplinary team of geneticists, genetic counselors, ethicists, psychologists, and survey design researchers. The questionnaires assessed sociodemographic characteristics, personal and family health information, prior use of genetic testing, motivations and concerns, psychological impact, risk perceptions, perceived utility and harms, and behavioral and medical actions. The questionnaire items were adapted from the Impact of Personal Genomics (PGen) Study[19], the MedSeq Project[6, 20–22], HealthSeq project[23] and CSER/CSER2 projects, and incorporated published measures of genetics knowledge[24], decision regret[25], anxiety[26], depression[27], intolerance of uncertainty[28] and attitudes towards genetic testing[29]. Specific items are described in more detail in the Supplemental Methods.

To increase clarity, the wording of questionnaire items was tailored to each collaborating site, including using site-specific names or only showing relevant response options. The questionnaire was expanded over time to facilitate comparisons with other studies and respond to changes in how EGS was offered and implemented. The later versions, administered to a subset of participants, incorporated or adapted items from a measure of health literacy[30], the Psychological Adaptation to Genetic Information Scale[31], a personal utility scale[32], the Perception of Uncertainties in Genome Sequencing (PUGS) scale[33], and the Feelings About genomiC Testing Results (FACToR) questionnaire[34].

### Participant recruitment and data collection

Through each collaborating site, the PeopleSeq Study recruited English speaking adults who had independently decided to pursue EGS (and when required by the collaborating site, had previously consented to be contacted about related research projects). Depending on the workflow and timing at the collaborating site, some sites initially discussed the PeopleSeq Study during in-person appointments, some sites sent their own personalized email invitations, some sites provided e-mails of eligible participants to the third party data collection firm, SoundRocket (Ann Arbor, MI), and some advertised the study via e-mail, flyers or websites. Participants who responded to the questionnaire via these passive entry points were asked additional questions to confirm eligibility and select the correct data collection pipeline (e.g., catch-up). If the initial survey invitations were sent by the data collection firm, participants received four reminders. Enrolled participants were entered into a random drawing to win an Apple Watch or a $50 Amazon gift card.

SoundRocket administered all questionnaires via a secure web-based platform and stored any identifying information collected through the questionnaires in a secured database accessible only to those approved. Regardless of study entry point, electronic consent was obtained from all participants prior to accessing the survey. All questionnaire items were optional and participants could freely navigate within the questionnaire. Questionnaires could be partially completed and could be completed in multiple sessions.

PeopleSeq investigators did not automatically receive the clinical testing report from the collaborating site. Depending on the site, at the end of the questionnaire participants were given the opportunity to share their clinical test report either by consenting for the site/project to directly send their report to SoundRocket, providing their public report identifier (PGP) or uploading an electronic copy of their report directly within the data collection system. SoundRocket redacted any identifying information in the report and then sent the de-identified genetic test report to the PeopleSeq Study team where it could be linked with the questionnaire data using the participant’s PeopleSeq study identifier. PeopleSeq did not receive any clinical records for participants; all clinical actions are self-reported by the participants.

### Data analyses

The analyses reported here were limited to participants who responded to the post-disclosure or catch-up questionnaires. Non-responder analysis for two cohorts with individual level non-responder demographic data available was previously reported[18]. For participants who completed the pre- and post-disclosure questionnaires (as opposed to the catch-up questionnaires), sociodemographic characteristics and motivations were reported in the pre-disclosure questionnaire; all other items were reported in the post-disclosure questionnaire. Previous work using a subset of this dataset (*n*=543) did not show statistically significant differences in outcomes, perceived utility and attitudes between the post-disclosure and catch-up questionnaires and so the results are presented here for both questionnaires combined[18].

Participants could report up to 3 results as free text items they self-assessed as important to them (in response to the questionnaire items “Did you receive any results that you felt were important to you” or “which results do you feel were most important to you”), termed “participant important results” (PIR). Multiple researchers coded these responses with regards to topic and directionality (i.e., “positive”, the presence of genetic findings, and “negative”, the absence of findings). The initial coding was reviewed by two researchers and differences resolved through consultation.

Multiple researchers reviewed all laboratory reports submitted by participants to identify individuals who received monogenic findings, termed “reported important results” (RIR). RIRs were not self-assessed by the participants. Here we defined monogenic findings as: heterozygous variants reported to be pathogenic or likely pathogenic (P/LP) for dominant conditions, or homozygous/compound heterozygous variants reported to be P/LP for recessive conditions. Monogenic findings were based on the original report received by the participant, not the current consensus interpretation for a variant. All monogenic findings were then further classified as “clinically important”, “possibly clinically important” or “not important” in the context of the participant’s reported medical history (as reported in survey responses and the provided laboratory report), current classifications for the variant as reported in ClinVar and other sources, the current ACMG guidelines for reporting secondary findings[35] and other professional guidelines, e.g., National Comprehensive Cancer Network.

All data analyses were performed using the R statistical package, version 4.3. Chi-square tests were used to evaluate differences by motivation for categorical variables.

## Results

### Cohort

Participants were recruited between October 2014 and December 2021. See Figure 1 for a flowchart. 6,431 participants were eligible for and individually invited to the study, however, because not all recruitment was individualized and trackable, the total number eligible individuals exposed to PeopleSeq recruiting information was larger. We estimate the number of eligible and invited participants to be 6,750. A total of 1,575 individuals consented to participate and enrolled in the study for an approximate participation rate of 23.3%; 1,147 individuals responded to a questionnaire after they received their genetic results (the data presented here) for a response rate of 72.8% among enrolled participants, and an approximate participation rate of 17.0% among all eligible and invited participants.

Table 1 summarizes the sociodemographic characteristics of participants who completed the PeopleSeq surveys. Supplemental Table 1 describes the testing provided by each project. The fractions of industry, academic, clinical and 3rd-party recruitments were 60.0%, 23.1% 16.2% and <1%, respectively. Most participants self-identified as white (87.4%), almost all had at least a college degree (91.5%) and most participants reported household incomes of $100,000 or more (78.9%). A fifth of participants (20.4%) reported that they were, or at the time of the questionnaire were studying to be, healthcare professionals. Almost half of participants (43.5%) reported undergoing some form of genetic testing prior to EGS. Of those who had previous genetic testing, a majority (57.1%) reported undergoing the previous testing because they were “curious about health and traits predicted by my genetic make-up”. A subset of participants were asked about the type(s) of genetic testing they underwent before doing EGS, and the majority reported direct-to-consumer (DTC) ancestry testing (57.3%) and/or DTC health-related testing (51.5%).

**Table 1:** Characteristics of participants (*n*=1147) who completed the post-disclosure or catch-up questionnaires.

| Characteristic | N (%)<sup>a</sup> |
| --- | --- |
| Age, mean (SD; range) in years | 52.7 (13.1; 21-91) |
| Gender |  |
| Female | 556 (48.5) |
| Male | 581 (50.7) |
| Other | 4 (0.3) |
| Race |  |
| Black or African American | 18 (1.6) |
| Asian | 43 (3.7) |
| White or Caucasian | 1003 (87.4) |
| More than one race or other race | 74 (6.5) |
| Hispanic or Latino | 33 (2.9) |
| Education |  |
| Less than college degree | 98 (8.5) |
| College degree | 265 (23.1) |
| Some graduate school | 436 (38.0) |
| Doctoral or professional degree | 340 (29.6) |
| Household annual income |  |
| <\$40,000 | 54 (4.7) |
| \$40,000-\$99,999 | 157 (13.7) |
| ≥\$100,000 | 905 (78.9) |
| Has biological children | 798 (69.6) |
| US resident | 928 (80.9) |
| Self-reported health |  |
| Excellent | 309 (26.9) |
| Very good | 481 (41.9) |
| Good | 207 (18.0) |
| Fair | 50 (4.4) |
| Poor | 8 (0.7) |
| Underwent prior genetic testing | 499 (43.5) |
| Provided report | 223 (19.4) |
| Project |  |
| Illumina Understand Your Genome (UYG)<sup>b</sup> | 381 (33.2) |
| Invitae | 307 (26.8) |
| Harvard Personal Genome Project (PGP) | 163 (14.2) |
| MedCan | 122 (10.6) |
| Mayo Clinic | 42 (3.7) |
| Genos | 36 (3.1) |
| Baylor (CEO, MD/PhD and Young Presidents' projects) | 35 (3.1) |
| Mount Sinai HealthSeq Project | 19 (1.7) |
| Genome Medical | 15 (1.3) |
| Other (LMM, MedSeq, Hudson Alpha, Veritas, 23andMe Exome Pilot, Other study or product)<sup>c</sup> | 27 (2.4) |
| Sequencing Technology |  |
| Genome Sequencing (GS) | 628 (54.8) |
| Exome Sequencing (ES) | 73 (6.4) |
| Targeted Sequencing (TS) | 442 (38.5) |

<sup>a</sup>Percentages may not sum to 100 due to missing responses or rounding
<sup>b</sup>Includes University of Vermont site
<sup>c</sup>For privacy, projects with fewer than 10 participants are combined

### Monogenic disease risk reports

A total of 223 individuals (19.4%) shared their EGS report with PeopleSeq researchers. From these reports we identified 37 individuals with 46 distinct monogenic findings, termed “reported important results” (RIR). Recurring findings (count in parentheses) included *HFE*:p.Cys282Tyr (4), *HFE*:p.His63Asp (5) (homozygous or compound heterozygous), *CHEK2*:p.Thr367fs (2) and *F5*:p.Arg534Gln (6).

Table 2 summarizes the self-reported actions of individuals who shared their report (see the Supplemental Methods for more detail). Individuals who received monogenic findings were more likely to report having received results they self-assessed as important to them (termed “participant important results” or PIR), discussing their results with a healthcare provider (HCP), and having tests, medical exams or procedures as a result of undergoing EGS. They also reported more medical visits on average than those who did not have a RIR.

**Table 2:** Actions for participants who provided their EGS test report who did not or did receive a monogenic finding (a “reported important result” or RIR). Monogenic findings were classified by PeopleSeq researchers based on the classification in the original report received by the participant.

| **Received monogenic findings (RIR)** | **No (n=186)** | **Yes (n=37)** |
| --- | --- | --- |
| Received information they felt was important | 53.8% | 83.8% |
| Discussed results with anyone | 81.7% | 94.6% |
| Discussed results specifically with family | 79.0% | 91.9% |
| Discussed results specifically with HCP | 38.9% | 67.6% |
| 1+ medical visits | 16.1% | 43.3% |
| Fraction that had tests, medical exams or procedures | 9.3% | 43.3% |
| Changed medications | 6.5% | 16.2% |
| Mean (SD) number of medical visits as a result of EGS | 0.3 (0.8) | 1.4 (3.4) |

Thirteen of the 46 RIR monogenic disease risk findings were classified as clinically important by PeopleSeq researchers (Supplemental Table 2) using the procedure described in the Methods. Two of the cases were previously reported by the Harvard Personal Genome Project (PGP)[36]: one determined to be not pathogenic by literature review and follow-up clinical evaluation and thus classified as not clinically important in this reinterpretation (*SCN5A*:p.Gly615Glu) and the second determined to be pathogenic based on follow-up clinical evaluation and thus classified as clinically important (*JAK2*:p.Val617Phe). Eleven of the thirteen findings classified were also reported as a PIR, i.e., self-assessed as important by the participant. The two clinically important RIR findings not reported as a PIR were a *HFE*:p.Cys282Tyr homozygote and a *BRCA2*:p.Lys3084Asnfs*26 heterozygote. However, these participants had already acted upon, or stated that they would soon act upon, these results in the future (as inferred from responses to other questionnaire items), suggesting they did indeed perceive the results as clinically important.

The first of the above two participants reported undergoing predispositional WES prior to the WGS testing that was the focus of this survey. In a subsequent follow-up questionnaire they reported undergoing a “serum ferritin test” in response to the homozygous *HFE*:p.Cys282Tyr finding, but because of prior WES, not the WGS testing. The second participant completed the PeopleSeq questionnaire in July 2015, but the *BRCA2* variant was only posted to their online report in May 2015. Thus it is possible that they were not aware of the result at the time they completed the PeopleSeq questionnaire. This individual completed a follow-up questionnaire in 2017. At that time they reported “*BRCA2*” as an important result and corresponding clinical follow-up care.

Two other participants with a clinically important RIR reported that they did not consult with an HCP other than the site team. One individual with a homozygous *SLC3A1*:p.Met467Thr finding associated with Cystinuria reported that they had already been diagnosed with kidney issues via other “medical test/procedures only”. One individual with a heterozygous *MSH2*:c.792+1del associated with Lynch syndrome reported that they had not consulted an HCP because “I do not have a physician at this time” but had discussed their results with their children (including information about the results and recommendations for testing). They reported their most important motivation to undergo testing was a confirmed genetic medical condition in their family of “Lynch syndrome”. The remaining participants who received clinically important RIR findings reported discussing their results with a primary care physician along with one or more of a “genetics specialist (e.g., genetic counselor or clinical geneticist)”, “cardiologist”, “oncologist”, “pediatrician” and “other specialist/healthcare provider”.

### Perception of important genetic results

A majority of participants (60.3%) reported receiving results that they self-assessed as important to them, termed “participant important results” (PIRs). Investigators coded the free text participant responses as “positive”, e.g., the presence of genetic findings, and “negative”, e.g. the absence of findings. Directionality could be inferred for 51.1% of PIRs, with approximately equal fractions positive and negative (24.5% and 26.5% of total, respectively).

A subset of participants (19.4%) provided their original laboratory or clinical report, enabling a comparison of actual versus self-reported findings. For individuals who provided their original report, we could link 91.4% of positive PIRs to specific variants or a set of consistent variants in their report (e.g., carrier variants) and 88.6% of negative PIRs were consistent with the report (i.e., no contradictory findings were present in the laboratory/clinical report). The remaining 11.4% of individuals with negative PIRs received findings annotated as clinically significant or P/LP in their laboratory or clinical reports.

Figure 2A shows the actions of individuals who reported different categories of PIRs. We observe that while those individuals who reported any negative PIRs indicated they discussed their results with family at similar rates as those who reported positive PIRs, they reported taking clinical actions, such as consulting an HCP, at lower rates. Figure 2B shows the fraction of participants who reported different categories of PIRs who also reported their genetic results were useful. We observe that individuals who reported negative PIRs also reported their results “reassure me that I am healthy” and “[gave] me information about specific diseases that I am concerned about” at higher rates.

**Figure 2:**
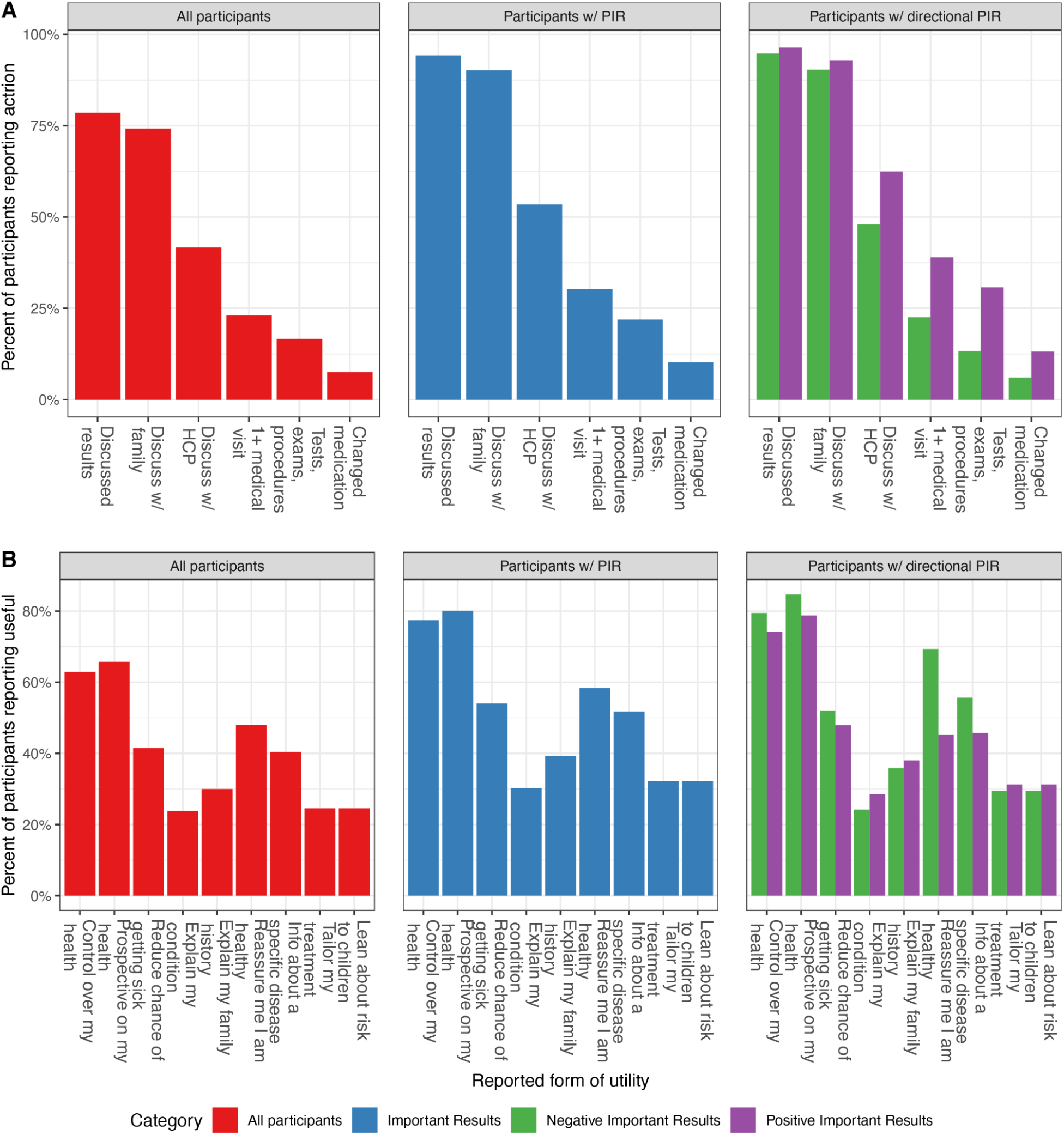
Actions and utility self-reported by participants post-disclosure and associated with self-assessed important results (PIR). Left-to-right in each panel: actions and utility for all participants, for participants who reported a PIR, and for participants who reported a PIR with any inferred directionality. Directional PIRs show participants who reported a positive vs. a negative PIR. A) Percentage of participants who reported action post-disclosure. B) Percentage of participants who agreed their results were useful.

We observed that participants who endorsed motivations related to a possible medical condition in their family (“there is a medical condition in my family that may be genetic”) as very or somewhat important, were significantly more likely to have reported a PIR than those who endorsed that motivation as “not” important or not applicable (Supplemental Table 3). Those same participants (who endorsed motivations related to family history) also reported negative PIRs at a higher rate, although the difference did not reach statistical significance (Supplemental Table 4).

### Reported post-disclosure actions

Figure 2A (left panel) shows participants’ self-reported actions after receiving their results. A majority of participants (78.5%) reported discussing their results with another person in some way, with most reporting discussing their results specifically with family members (74.1%).

Smaller fractions reported discussing their results with an HCP other than the site team (41.7%), having 1 or more medical visits (23.1%), undergoing tests, medical exams or procedures related to their results (16.7%) and/or making changes to their use of prescription and non-prescription medications, supplements, or use of alternative medicine (7.6%) because of their genetic test results. Figure 3A shows the specialties of the HCPs that participants reported consulting about their results. Primary care providers were the most common to be consulted (32.3% of individuals who reported consulting an HCP). Figure 3B shows which family members participants reported discussing their results with. Figure 3C shows which medical tests/procedures participants reported undergoing because of their results.

**Figure 3:**
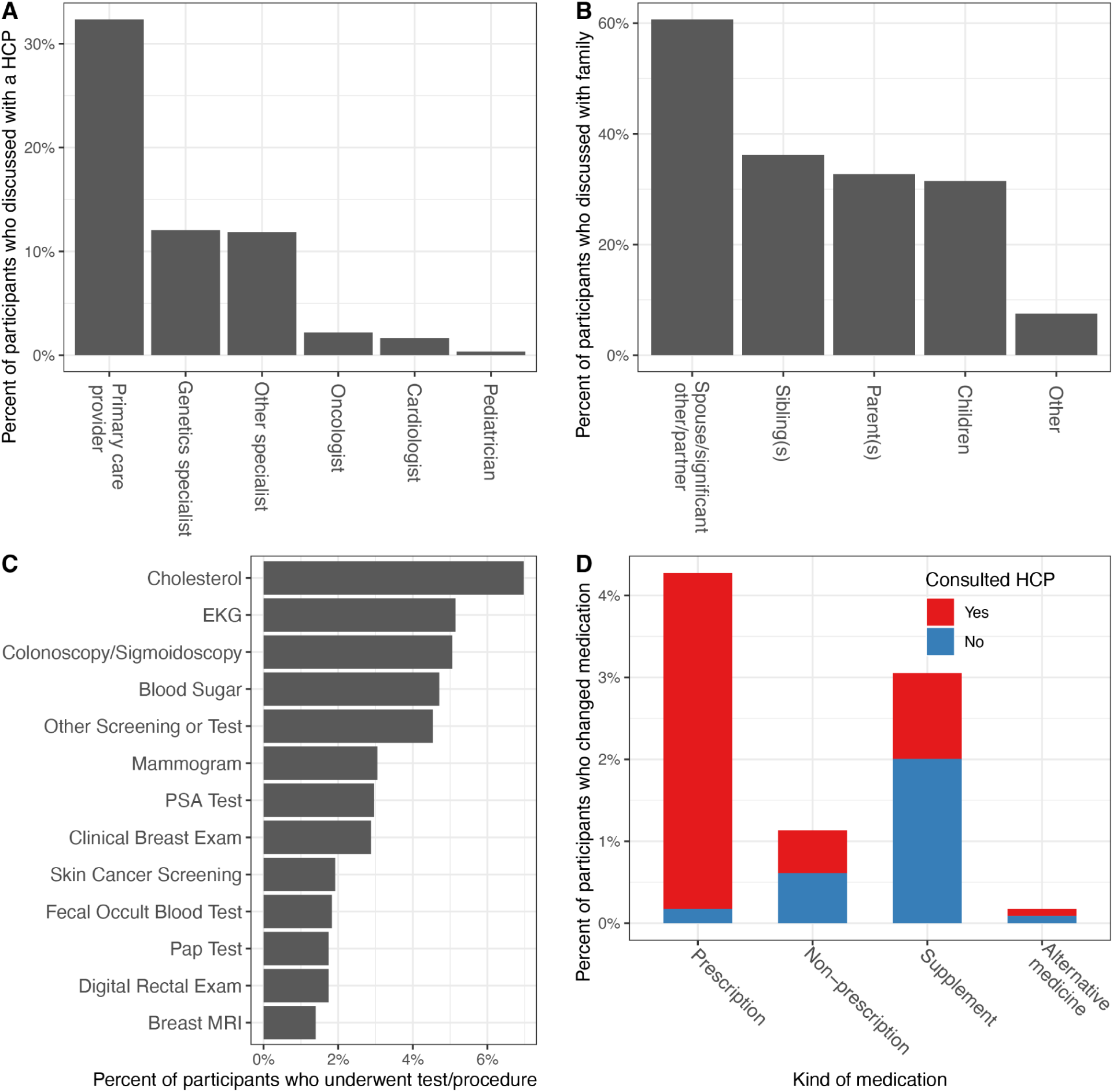
Detail of actions reported by participants post-disclosure. The percent of participants who reported discussing their results, 1+ medical visits, undergoing tests, exams or procedures, and/or making a change to their medications as result of their genomic testing are shown in Figure 2A (left panel). A) Of participants who reported discussing their results with any HCP, percent who reported discussing with a specific type of HCP. B) Of participants who reported discussing their results with family, percent who reported discussing their results with specific family members. C) Of participants who reported undergoing tests, exams or procedures, percent of participants who reported a specific type of test, exam or procedure. D) Of participants who reported changing their medication, percent who reported changing a specific type of medication and whether they consulted a HCP prior to making the change.

Figure 3D shows which types of medications individuals reported changing in response to their EGS results and whether they reported consulting their HCP prior to making that change. Only 2 individuals reported making a change to their prescription medications without consulting an HCP. Those who reported making a change to their medication(s) reported making a mean (SD) of 1.86 (1.66) such changes. Participants’ motivations for these medication changes included indications of disease (e.g., cholesterol levels), or genetically informed predictions of medication effectiveness, dosing and side effects. Multiple participants described their genetic testing providing specific evidence to confirm personal observations and effect change in clinical management. For example, one participant wrote: “*CYP2D6* poor metabolizers do not get sufficient analgesia from codeine, hydrocodone, oxycodone, or tramadol, which has always been a problem for me. [Cohort] results enabled me to request different pain options and have data to back it up.” Another participant noted “Family history had to be established in order for my insurance carrier to pay for Repatha infusion.”

### Motivations for EGS

Figure 4 shows the fraction of participants who endorsed each of a potential set of motivations for pursuing EGS (presented as 3 options: not at all, somewhat, or very important). Almost all respondents reported “interest in finding out about my personal disease risk” (91.4%) and “curiosity about my genetic makeup” (90.7%) as a very or somewhat important motivation.

**Figure 4:**
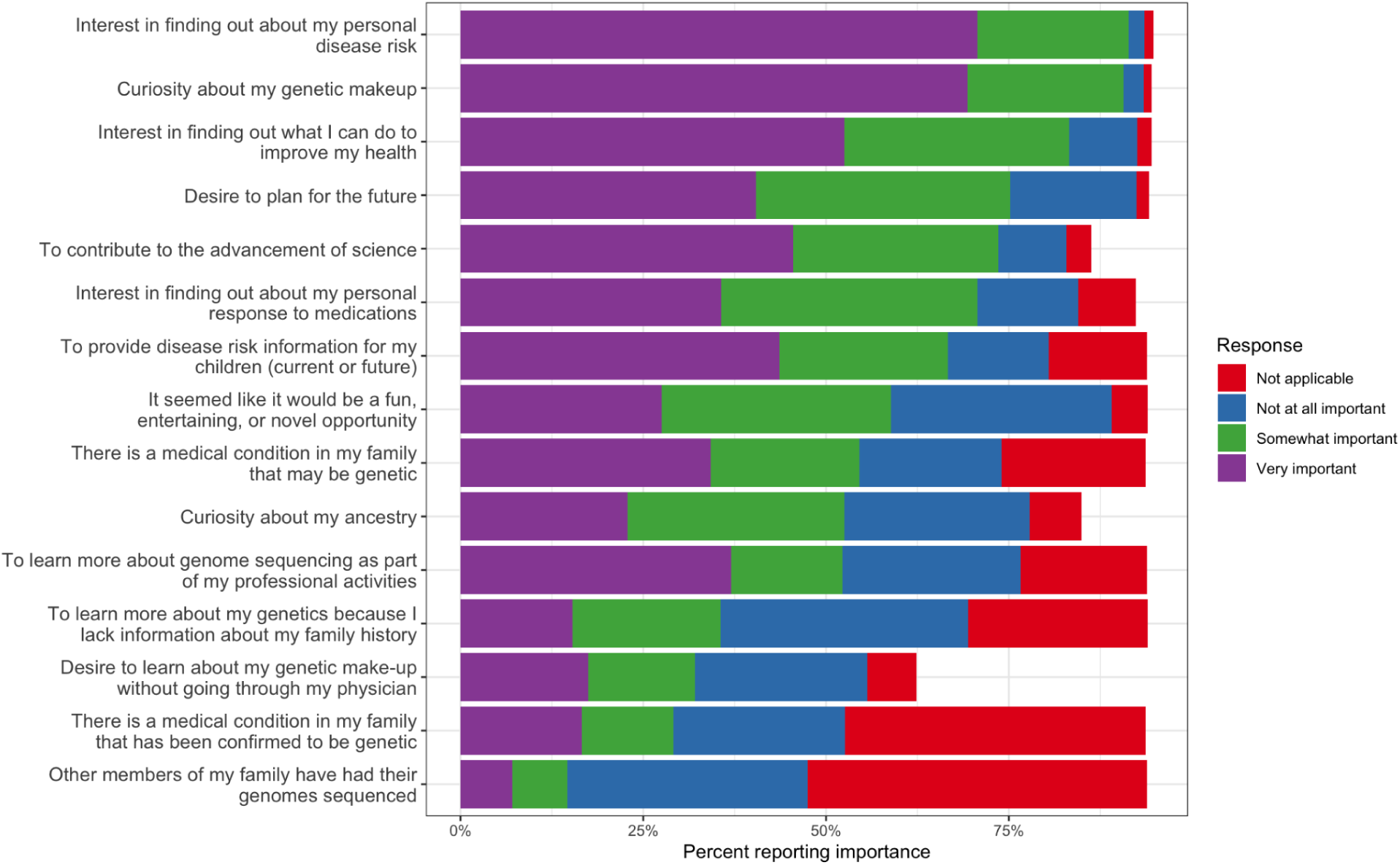
Percentage of respondents reporting the importance of different factors in deciding to pursue EGS. Motivations ordered by the sum of “Very important” and “Somewhat important” responses.

Additionally more than half of participants reported a quasi-indication-based motivation for pursuing EGS, indicating “there is a medical condition that runs in my family that may be genetic” (54.6%) as a very or somewhat important motivation. During data collection, the questionnaire was modified to also ask the most important motivation (Supplemental Table 5). Out of the subset of participants who received this question (*n*=1062), 27.8%, 11% and 5.5%, respectively, reported the motivations above as their most important motivation.

### Illustrative examples

We observed illustrative examples of the proposed benefits and concerns for EGS. One individual reported a “*MSH2* mutation” identified by EGS, leading to a diagnosis based on their EGS results, medical/family history and medical tests/procedures. In response to why they felt the personal genetic sequencing was/was not valuable, they responded that they “Identified a hereditary issue that myself and others did not know about and could have potentially died from.”. An individual reported distress in response to “Cardiomyopathy risk”. In response to why they feel the personal genetic sequencing was/was not valuable, they responded “I had an unexpected result that caused me a lot of anxiety and took a long time to resolve. It ended up probably not being a big worry after all but there is still some uncertainty” and in summary they stated “I ended [up] having a bad experience but I don’t regret going through the testing”. A different individual made statements that suggested the possibility of false reassurance about their breast cancer risk, reporting “Probability of breast cancer is low (good, because my mother died of it)” as a self-assessed important result (a PIR). They described their risk as “much lower” than the average person. One respondent may exemplify overmedicalization as they reported seeking follow-up care for uncertain findings and variants not found in their report.

More than half of participants reported as somewhat or very important the quasi-indication-based motivation “Concern about a medical condition that runs in my family that may be genetic” (54.6%), indicating that some participants were seeking explanatory as opposed to predictive results. Two illustrative examples were individuals whose primary motivation was establishing a familial hypercholesterolemia diagnosis to get approval for Repatha: “Statins were giving me kidney disease, so [I] had to make a change. FHC diagnosis was instrumental in get[ting] me approved for Repatha.” and “Required to establish Hypercholesterolemia treatment with Repatha.” Other individuals reported the testing helped explain family history. For example an individual with a pathogenic *BRCA2* finding described the family history that motivated their testing: “My mother had breast cancer, 6 months later my aunt (mother’s sister) dx with breast cancer. Nobody had genetic testing”. In some cases, like the first two above, the genetic testing was obtained in consultation with their HCP(s). For others, the testing seemed to be prompted by prior unsatisfactory clinical interactions. One described why their personal genetic testing was valuable: “Supports my clinical experience. I am NOT a drug seeker. Opioids just don’t work well for me. Now that this info is in my chart, I expect that I will receive an effective dose should I need pain Rx in the future without judgment or hassle.”

## Discussion

Here we presented data from the PeopleSeq Study, a collaborative effort between 18 academic, clinical and commercial EGS projects/laboratories to collect data on the impacts of this testing across the different and diverse contexts in which EGS is in use. Consistent with previous reports of both clinical benefits and potential harms[37], we observed examples of individuals learning about previously unknown disease risks, and instances of concerns, including psychological distress, false reassurance and potentially unnecessary follow-up care. In aggregate, however, we observed that individuals’ self-reported important results were consistent with actual reported findings (available for 19.4% of participants), and that individuals with findings most likely to be relevant to their health (e.g., related to ACMG secondary findings and/or ClinGen actionability genes) were also more likely to report discussing their results with an HCP and undergoing follow-up care. Among individuals who provided test reports and who received clinically important monogenic findings, all either directly described those findings as important and/or reported acting on those results. Most of these participants (11/13) reported discussing their results with one or more HCP(s). Those individuals who reported self-assessed important genetic findings (PIRs) inferred to be “negative” (i.e., the absence of findings) were equally likely as those reporting self-assessed important findings inferred to be “positive” to have discussed their results with a family member, but less likely to have discussed their results with a HCP or undergone follow-up care.

While 41.7% of participants reported discussing their results with their HCP (other than the site team), only 23.1% reported having one or more subsequent medical visits as a direct result of their EGS testing. This suggests that individuals are integrating these discussions into their regular care rather than scheduling separate interactions. It may also suggest that the results were not considered medically actionable by the HCP. Expanding on previous results[18], the fraction of individuals discussing their results with a HCP is higher in this study than reported for DTC genotyping-based testing[38]. That difference may reflect the different role for clinicians in ordering EGS testing in many of the collaborating projects, as well as the scope and nature of the results generated by sequencing vs. genotyping.

Previous studies have suggested that when a large number of defined medically relevant genes are included in the interpretive pipeline, 10-20% of EGS participants will receive a P/LP variant for a monogenic disorder, with a larger fraction receiving pharmacogenomic, carrier or polygenic disease risk findings (if offered)[13–17, 39–42]. The range of findings reflects differing target populations, scope of testing (targeted panels of different sizes vs. exome vs. genome) and reporting thresholds (e.g., specific gene list vs. all known pathogenic variants). We observed that a similar fraction of participants received P/LP variant findings (among participants who provided their report).

A majority of individuals (60.3%) reported receiving PIRs, results they self-assessed as important to them; of those, approximately equal fractions could be inferred as “positive” (the presence of a variant) and “negative” (the absence of a variant) findings. The high rate of self-described important results is consistent with the larger fraction of individuals who received carrier and/or pharmacogenomic findings and a broader definition of personal utility for personal genetic testing than just receiving P/LP monogenic disease-related findings[32, 43, 44]. The difference between the rate of self-assessed important results and the rate at which participants reported discussing their results with an HCP other than the site team may also reflect the direct role of clinicians in the EGS testing process at collaborating projects.

Participants expressed a variety of motivations for undergoing EGS, notably both general curiosity and specific interest in their personal disease risk. More than half of PeopleSeq participants (54.6%) also reported a quasi-indication-based motivation of “Concern about a medical condition that runs in my family that may be genetic”. This quasi-indication-based usage is consistent with previous observations that individuals may be motivated to undergo EGS due to an actual or perceived risk for a genetic condition included in the test[45]. These individuals may be using EGS as a quasi-diagnostic test or in lieu of other forms of indication-based testing[46]. Similar motivations were reported in the context of DTC genotyping, where users expressed interest in findings related to their family history and existing medical conditions[47, 48]. Individuals who reported a family history-related motivation were significantly more likely to report receiving PIRs than those who did not. Previous comparisons of diagnostic and elective (EGS) patients showed overlapping motivations between those groups, with a “sizable minority” of elective patients interested in diagnostic information[46]. A subset of individuals reported pursuing EGS for explicitly diagnostic purposes, including examples of individuals seeking to establish a familial hypercholesterolemia diagnosis in consultation with their HCPs, and individuals seeking to confirm personal observations and effect change after previously unsatisfactory clinical interactions.

The range of motivations suggest that these EGS participants are from several distinct groups, including both unselected individuals undergoing screening out of general interest and individuals who self-selected due to actual or perceived genetic disease risk. As a result of the latter group, EGS cohorts may be enriched for individuals at increased risk for a genetic condition. Such demographics would be consistent with the increased rates of positive findings reported in EGS cohorts compared to unselected populations[16, 49], and could have implications for pre- and post-test counseling, particularly related to the interpretation of negative results, or more precisely in the context of EGS, the absence of a positive result.

Consistent with prior interview-based studies[50], PeopleSeq EGS participants considered negative results important. But the negative predictive value of those results will depend on the design of the specific test (which is varied and evolving) and the individual’s personal and family history. Even if precisely defined, the interpretation of those results may depend on an individual’s genetic literacy, risk perception, cultural background and other factors[51].

For some individuals, EGS may have provided a means to overcome barriers to obtaining quasi-indication-based or even indication-based diagnostic testing, such as cost, access to HCPs who can provide such testing, and limited integration of genetic testing into existing healthcare delivery systems[52, 53]. However, EGS tests are not designed for individuals with a personal or family history of a specific medical condition, and are typically not an equivalent alternative to the corresponding diagnostic test. These results reinforce the need for a comprehensive regulatory framework and accompanying practice guidelines for EGS that address the different ways individuals use this technology[54, 3, 37, 53]. Two such resources are the 2021 ACMG’s “points to consider”[3] and the 2023 practice resource published by the National Society for Genetic Counselors[37].

There are limitations to this study. Eligibility was restricted to English-speaking adults. The sample size of *n*=1147 incorporated a range of testing contexts. However, due to the distributed project structure and opportunistic recruiting approach, the overall participation rate was approximately 17.0%. Those who did participate may be different from the population undergoing EGS (non-responder bias) and different from the general public. Responders may have been interested in genomics (personally or professionally) or have had a more positive experience than non-responders. Previous non-responder analyses in early PeopleSeq cohorts showed that while demographic differences between responders and non-responders were statistically significant, they were similar to differences reported in other studies[18].

The participants in this study were early adopters of EGS, and like early adopters of other technologies, are not necessarily representative of the general population[55]. Participants sought out, and in some cases paid for, EGS and so may have been more likely to perceive it as useful and the results as important. Consistent with other studies of EGS[56] and DTC genetic testing early adopters, PeopleSeq participants are generally white with high educational attainment and high household incomes. Due to the upstream project recruitment strategies, PeopleSeq participants are enriched for current or future health professionals. Their results may not generalize to other populations. However, since little is currently known about the outcomes for individuals undergoing EGS; studying those individuals who are actually using these technologies can provide useful data to inform our understanding of and approach to EGS in the future. Data from health professionals who underwent EGS can help understand how experiential learning could influence their professional practice[57].

Participants had heterogeneous testing experiences, including different contexts (e.g., clinical vs. research), role of clinical supervision, forms of pre- and post-test counseling, scale of sequencing (targeted vs. exome vs. genome) and reporting practices. The different scale of testing could influence the number and types of results returned, participants’ actions and their perception of the utility of EGS. Recruitment was conducted over a multi-year period in which the availability, context, costs and capabilities of EGS changed substantially. Genome interpretation has evolved over that time; variant findings that were previously reported as P/LP may be classified differently now[58, 59]. Since the focus is on participant behaviors and outcomes, we classified variants based on both the information the participant originally received and the current consensus interpretation.

Participants’ actions and their important results (PIRs) were self-reported, not directly measured[60]. The PeopleSeq Study had access to participant’s EGS reports for a subset of participants (19.4%) enabling comparison between actual and self-reported results. Self-reported important findings were generally consistent with the reports when available, indicating PIRs are informative for individuals’ actual results. However, since participants needed to explicitly opt-in to sharing the report they received and needed to provide an electronic copy of that report themselves (excluding PGP and Mount Sinai cohorts), the distribution of reports by cohort differed from the study as a whole. Results related to participant-provided reports may be impacted by non-responder bias.

Despite these limitations, and their potential impacts on generalizability, this study offers insight into motivations for and clinical actions of participants in EGS programs across a wide range of contexts. We observe that early adopters of EGS are motivated by general interest in their health as well as quasi-indication-based considerations such as family history. A majority of participants learned results they considered important and shared these results with family members, but a much smaller segment engaged HCPs with their results. These data can be used to inform future implementation of EGS programs.

## List of abbreviations

EGS: Elective genome sequencing
DTC: Direct-to-consumer
PIR: Participant important results
RIR: Reported important results
HCP: Healthcare provider

## Declarations

### Ethics approval and consent to participate

The Partners Healthcare Human Research Committee approved this study along with Baylor College of Medicine Institutional Review Board and Harvard Medical School Institutional Review Board for PGP. Informed consent was obtained electronically from all participants. Individual data was de-identified by the survey administrator prior to analysis. This study conforms to the Declaration of Helsinki.

### Consent for publication

Not applicable

### Availability of data and materials

The datasets used and/or analyzed during the current study are available from the corresponding author on reasonable request

## Author contributions

Conceptualization: R.C.G.; Data Curation: M.D.L., T.B., S.M.A.; Analysis: M.D.L., T.B., S.M.A., T.S. S.P.; Funding Acquisition: R.C.G.; Investigation: M.D.L., S.M.A., T.M.B., S.D.C., T.J.C., A.T.E., D.E.N., S.P., Project Administration: M.D.L., T.B., S.M.A, D.E.N..; Resources: M.D.L., T.M.B., J.L.A., S.D.C., T.J.C., E.D.E., A.T.E., T.S., H.A., G.C., E.V.H., M.H., L.N.K., P.K., D.L., T.M., M.M., V.P., E.E.S., B.G.T., B.Z., R.C.G.; Supervision: A.L.M., R.C.G.;, Writing-original draft: M.D.L.; All authors reviewed and approved the manuscript.

## Acknowledgements

Not applicable

## PeopleSeq Team Members

Bethany Zettler, Paul Kruszka, Jane Juusola, Kirsty McWalter, Steven Bleyl, Ed Esplin, Eden Haverfield, Tara Schmidlen, Lazaridis Konstantinos, Jennifer Anderson, Heather Andrighetti, Allison Hazell, Jessica Gu, Madhuri Hegde, Jennifer Hogan, Vaibhav Pandya, George Church, Madeleine Ball, Thomas May, Molly McGinnis, Eric Schadt, Bastian Greshake Tzovaras, Debra Leonard, Robert C. Green, Michael Linderman, Amy McGuire, Kurt Christensen, Daiva E Nielsen, Thomas Caskey, Stacey Pereira, Emilie Zoltick, Tala Berro, Sophia Adelson, Scott Crawford, Robert Young; Jillian Hunsanger; Sophia Bradley, Tshaka Cunningham, Altovise Ewing-Crawford, Scott Roberts, Luisel Ricks-Santi, James Lillard Jr., Kareem Washington, Anthony Johnson

