## Supplemental Methods, Tables and Figures for "Elective genomic sequencing for adults in research, clinical and commercial contexts"

#### Supplemental Methods, Tables and Figures

Michael D. Linderman[1]*, Sophia M. Adelson[2,3], Tala M. Berro[2], Jennifer L. Anderson[4], Scott D. Crawford[5], Tshaka J. Cunningham[6], Edward D. Esplin[7], Altovise T. Ewing-Crawford[8], Daiva E. Nielsen[9], Stacey Pereira[10], Tara Schmidlen[11], Heather Andrighetti[12], Steven Bleyl[13], George M. Church[14,15,16,17], Eden V Haverfield[18], Madhuri Hegde[19], Lazaridis N. Konstantinos[4], Paul Kruszka[20], Debra Leonard[21], Thomas May[22], Molly McGinniss[23], Vaibhav Pandya, Eric E. Schadt[24], Bastian Greshake Tzovaras[14], Bethany Zettler[2], Amy L. McGuire[10], Robert C. Green[2,25,26,27]* and the PeopleSeq Study Team

[1]Department of Computer Science, Middlebury College, Middlebury, VT, USA
[2]Department of Medicine, Mass General Brigham, Boston, MA, USA
[3]Stanford School of Medicine, Stanford, CA, USA
[4]Center for Individualized Medicine, Mayo Clinic. Rochester, MN, USA
[5]Sound Rocket, Ann Arbor, MI, USA
[6]Faith Based Genetic Research Institute, Detroit, MI, USA
[7]Labcorp Genetics, San Francisco, CA, USA
[8]InGENEuity, Pulaski, TN, USA
[9]School of Human Nutrition, McGill University, Ste. Anne-de-Bellevue, QC, Canada
[10]Center for Medical Ethics & Health Policy, Baylor College of Medicine, Houston, TX, USA
[11]Nest Genomics, Beaverton, OR, USA
[12]Medcan, Toronto, ON, Canada
[13]Genome Medical Services, S. San Francisco, CA, USA
[14]Open Humans Foundation, Sanford, NC, USA
[15]Harvard Personal Genome Project, Harvard Medical School, Boston, MA, USA
[16]Department of Genetics, Harvard Medical School, Boston, MA, USA
[17]Wyss Institute for Biologically Inspired Engineering, Harvard University, Boston, MA, USA
[18]Genomic Life, La Jolla, CA, USA
[19]Revvity, Waltham, MA, USA
[20]Pediatric Genetics, University of Virginia, Charlottesville, VA, USA
[21]Department of Pathology and Laboratory Medicine, University of Vermont, Burlington, VT, USA
[22]Department of Community and Behavioral Health, Elson S. Floyd College of Medicine, Washington State University, Spokane, WA, USA
[23]Alcance Genomics, San Diego, CA, USA
[24]Department of Genetics and Genomic Sciences, Icahn School of Medicine at Mount Sinai, New York, NY, USA

[25]Ariadne Labs, Boston, MA, USA
[26]Broad Institute of Harvard and MIT, Cambridge, MA, USA
[27]Harvard Medical School, Boston, MA, USA

* Corresponding Author

### Supplemental Methods

**Description of self-reported actions**

The determination of post-disclosure actions are described in more detail below:

*Participant important results (PIR)*: Importance was self-assessed and in response to the questionnaire items “Did you receive any results that you felt were important to you” or “which results do you feel were most important to you”. Participants could report up to 3 such results as free text items.

*Discussed results (with anyone)*: Derived from an initial item “Have you discussed your results with anyone” followed by a select-all-that-apply item for different subgroups if the answer was “Yes” , or the union of all subgroups when the select-all item was administered to all.

*Discuss w/ family* and *Discuss w/ HCP*: Participants specifically reported having discussed their results with “Family member(s) (including spouse / significant other / partner)” and “Your healthcare provider (other than the [site] team)”.

*1+ medical visits*: Participants self-reported 1 or more “medical visits with a physician or other healthcare provider (other than the [site] team) have you had as a direct result of receiving your [site] results (in other words, visits you would not have had if you had not had personal genome sequencing)”.

*Tests, medical exams, or procedures*: Derived from an item “As a result of seeing your [site] results, have you had any tests, medical exams, or procedures?” or responding “Yes” to any of a set of tests or exams the participant “had because of your [site] results or follow-up care prompted by your [site] results?”.

*Changed medications*: Participants self-reporting “Because of your [site] results, have you made any changes to your use of medications (including prescription and non-prescription), supplements, and/or use of alternative medicine?”.

### Supplemental Tables and Figures

Supplemental Table 1: Sites participating in the PeopleSeq consortium. Setting is one of Clinical, Academic, Industry or 3rd-party (where participants upload their own data). Type is one of targeted Panel, Exome and/or Genome sequencing. Kinds of results returned, approach to pre-test counseling and results disclosure and the availability of raw data shown for each site. Enrollment describes whether a site recruited participants to both pre-post surveys (PP) and catch-up (CU) or just CU.

| Site | Setting | Type | Results Returned | Pre-test counseling | Results disclosure | Raw data | Enrollment |
|---|---|---|---|---|---|---|---|
| Preventative Genomics Clinic (Brigham & Women's Hospital) | C | PEG | Test dependent | Genetic counselor | Ordering HCP (MD/GC) | Upon Request | CU |
| Baylor College of Medicine YPO, CEO and MD/PhD | A | E | Monogenic disease findings | Investigator (MD) | Investigator (MD) with non-clinical report | None | CU |
| GeneDx | I | E | Monogenic disease findings | Ordering HCP | Ordering HCP | Upon Request | PP/CU |
| Harvard Personal Genome Project (PGP) | A | G | Variants with interpretation and lit. annotations | None | Online with semi-automated non-clinical report | VCF | PP/CU |
| HudsonAlpha Institute for Biotechnology | A | G | Monogenic disease findings | Genetic counselor | Investigator (MD/GC) with clinical report | None | CU |
| Illumina Understand Your Genome (UYG) | I | G | Monogenic disease findings, PGx | Ordering HCP | Online, clinical report sent to ordering HCP | VCF via HCP or by request | PP/CU |
| Invitae | I | P | Monogenic disease findings | Ordering HCP | Ordering HCP | Upon Request | CU |
| Mayo Clinic | C | PEG | Monogenic disease findings | Genetic counselor | Genetic Counselor | Upon Request | CU |
| Mount Sinai School of Medicine (MSSM) | A | G | Monogenic disease findings, PGx, polygenic risk | Investigator (GC) | Investigator (MD) with non-clinical report | BAM, VCF | CU |
| OpenSNP | 3rd | N/A | N/A | N/A | N/A | N/A | CU |
| PerkinElmer | I | G | Monogenic disease findings | Ordering HCP, Genome Medical | Ordering HCP, Genome Medical | Upon request | CU |
| University of Vermont (UVM) (via Illumina UYG) | A | G | Monogenic disease findings, PGx | Ordering HCP | Online, clinical report sent to ordering HCP | VCF via HCP or by request | PP/CU |
| Genome Medical | C | PEG | Test dependent | Genetic counselor | Ordering HCP (MD/GC) | Upon Request | PP/CU |
| MedSeq | A | G | Monogenic disease findings | PCP or Cardiologist | PCP or Cardiologist | Upon Request | CU |
| Medcan | C | P | Monogenic disease findings | Genetic counselor | Genetic Counselor | Upon Request | PP/CU |
| Takeda | I | G | Monogenic disease findings | Via Genome Medical | Via Genome Medical | Yes, to all | CU |
| Genos/NantOmics | I | E | 27 traits, "gene cards", variants with lit. annotations | Available for separate fee | Online | BAM, VCF | CU |

Supplemental Table 2: The subset of 46 monogenic variants in participant's EGS reports (RIR) further classified by PeopleSeq researchers as clinically important. Variant description and conditions are as described in the report received by the participant. If possible, variants were linked to results self-assessed by participants as important to them (PIR). The first two participants are discussed in the main text.

| **Variant as described in report** | | | | **Self-assessed as Important** |
|---|---|---|---|---|
| **Variant** | **Class** | **Zygosity** | **Condition(s)** | |
| HFE:c.845G>A (p.Cys282Tyr) | P | Homozygous | Hereditary Haemochromatosis | |
| BRCA2:c.9252_9255delAACAinsTT (p.Lys3084fs) | P | Heterozygous | Breast Cancer | |
| JAK2:c.1849G>T (p.Val617Phe) | P | Heterozygous | Myeloproliferative diseases | ✓ |
| SLC3A1:c.1400T>C (p.Met467Thr) | P | Homozygous | Cystinuria | ✓ |
| CHEK2:c.1100delC (p.Thr367fs) | P | Heterozygous | CHEK2-Related Cancer Susceptibility | ✓ |
| BRCA2:c.5857G>T (p.Glu1953Ter) | P | Heterozygous | BRCA2 Related Disorders | ✓ |
| FLNC Deletion (Exons 27-28) | P | Heterozygous | Neuromuscular and heart-related conditions | ✓ |
| GLA c.644A>G (p.Asn215Ser) | P | Hemizygous | Fabry Disease | ✓ |
| F5:c.1601G>A (p.Arg534Gln), F2:c.*97G>A | P | Heterozygous | Prothrombin-Related Thrombophilia, Factor V Leiden Thrombophilia | ✓ |
| HOXB13:c.251G>A (p.Gly84Glu) | Increased Risk Allele | Heterozygous | Prostate Cancer | ✓ |
| CHEK2:c.1100del (p.Thr367fs) | P | Heterozygous | Hereditary Cancer | ✓ |
| MSH2:c.792+1del | LP | Heterozygous | * | ✓ |
| BRCA2:c.5238dup (p.Asn1747Ter) | P | Heterozygous | * | ✓ |

* Report available to PeopleSeq project included interpretation but not the associated condition

Supplemental Table 3: Fraction of individuals endorsing specific motivations as very/somewhat important vs. not important/not applicable who reported receiving results important to them (PIR). $X^2$ test statistic, degrees of freedom and p-value are reported for each motivation.

| **Motivation** | **Very or somewhat important** | | **Not important or NA** | | |
|---|---|---|---|---|---|
| **Important Results (PIR)** | **Yes** | **Not sure** | **Yes** | **Not sure** | **$X^2$ test** |
| Curiosity about my genetic makeup | 0.676 | 0.153 | 0.698 | 0.116 | 0.4(2) p=0.801 |
| Interest in finding out about my personal disease risk | 0.677 | 0.154 | 0.622 | 0.081 | 4.8(2) p=0.0895 |
| Interest in finding out what I can do to improve my health | 0.677 | 0.162 | 0.664 | 0.080 | 10.6(2) p=0.00495 |
| Interest in finding out about my personal response to medications | 0.664 | 0.167 | 0.696 | 0.114 | 4.1(2) p=0.13 |
| Desire to plan for the future | 0.682 | 0.163 | 0.645 | 0.114 | 10.2(2) p=0.00623 |
| It seemed like it would be a fun, entertaining, or novel opportunity | 0.656 | 0.154 | 0.708 | 0.151 | 4.4(2) p=0.112 |
| Desire to learn about my genetic make-up without going through my physician | 0.629 | 0.180 | 0.690 | 0.176 | 4.4(2) p=0.112 |
| Other members of my family have had their genomes sequenced | 0.748 | 0.135 | 0.660 | 0.156 | 5.3(2) p=0.0699 |
| To learn more about my genetics because I lack information about my family history | 0.717 | 0.161 | 0.648 | 0.147 | 11.3(2) p=0.00347 |
| To provide disease risk information for my children (current or future) | 0.687 | 0.145 | 0.653 | 0.167 | 1.2(2) p=0.556 |
| There is a medical condition in my family that may be genetic | 0.725 | 0.135 | 0.605 | 0.177 | 16.8(2) p=2.27E-4 |
| There is a medical condition in my family that has been confirmed to be genetic | 0.708 | 0.144 | 0.660 | 0.156 | 2.7(2) p=0.266 |
| To learn more about genome sequencing as part of my professional activities | 0.675 | 0.138 | 0.676 | 0.171 | 3.5(2) p=0.174 |
| To contribute to the advancement of science | 0.646 | 0.171 | 0.723 | 0.131 | 3.1(2) p=0.218 |
| Curiosity about my ancestry | 0.623 | 0.191 | 0.693 | 0.134 | 6.1(2) p=0.0465 |

Supplemental Table 4: Fraction of individuals endorsing specific motivations as very/somewhat important vs. not important/not applicable who reported any important results (PIR) coded as negative. $X^2$ test statistic, degrees of freedom and p-value are reported for each motivation.

| **Motivation** | **Very or somewhat important** | **Not important or NA** | |
|---|---|---|---|
| **Negative important results (PIR)** | **Yes** | **Yes** | **$X^2$ test** |
| Curiosity about my genetic makeup | 0.229 | 0.227 | 0.0(1) p=1 |
| Interest in finding out about my personal disease risk | 0.230 | 0.179 | 0.3(1) p=0.587 |
| Interest in finding out what I can do to improve my health | 0.223 | 0.264 | 0.8(1) p=0.358 |
| Interest in finding out about my personal response to medications | 0.195 | 0.337 | 21.2(1) p=4.21e-06 |
| Desire to plan for the future | 0.230 | 0.229 | 0.0(1) p=1 |
| It seemed like it would be a fun, entertaining, or novel opportunity | 0.222 | 0.241 | 0.4(1) p=0.533 |
| Desire to learn about my genetic make-up without going through my physician | 0.212 | 0.248 | 1.1(1) p=0.293 |
| Other members of my family have had their genomes sequenced | 0.232 | 0.230 | 0.0(1) p=1 |
| To learn more about my genetics because I lack information about my family history | 0.213 | 0.240 | 0.9(1) p=0.342 |
| To provide disease risk information for my children (current or future) | 0.232 | 0.224 | 0.0(1) p=0.83 |
| There is a medical condition in my family that may be genetic | 0.249 | 0.205 | 2.6(1) p=0.104 |
| There is a medical condition in my family that has been confirmed to be genetic | 0.225 | 0.232 | 0.0(1) p=0.846 |
| To learn more about genome sequencing as part of my professional activities | 0.219 | 0.245 | 0.9(1) p=0.349 |
| To contribute to the advancement of science | 0.216 | 0.301 | 4.7(1) p=0.0299 |
| Curiosity about my ancestry | 0.194 | 0.280 | 9.0(1) p=0.00264 |

Supplemental Table 5: The fraction of individuals reporting specific motivations as the “most important”. This item was only included in a subset of surveys (*n*=1062). Fractions do not add to one due to missing data.

| **Motivation** | **Fraction** |
|---|---|
| Interest in finding out about my personal disease risk | 0.278 |
| Curiosity about my genetic make-up | 0.110 |
| To learn more about genome sequencing as part of my professional activities | 0.073 |
| To provide disease risk information for my children (current or future) | 0.063 |
| Other | 0.059 |
| Interest in finding out what I can do to improve my health | 0.056 |
| There is a medical condition in my family that may be genetic | 0.055 |
| To contribute to the advancement of science | 0.039 |
| Desire to plan for the future | 0.019 |
| There is a medical condition in my family that has been confirmed to be genetic | 0.017 |
| To learn more about my genetics because I lack information about my family history | 0.015 |
| Interest in finding out about my personal response to medications | 0.012 |
| It seemed like it would be a fun, entertaining, or novel opportunity | 0.015 |
| Curiosity about my ancestry | 0.00657 |
| Desire to learn about my genetic make-up without going through my physician | 0.006 |
| Other members of my family have had their genomes sequenced | 0.001 |
